# Transient frontal spectral events from EEG predict antidepressant response to sertraline in depression

**DOI:** 10.64898/2026.01.26.26344862

**Authors:** Darcy A. Waller, Linda L. Carpenter, Stephanie R. Jones

## Abstract

Resting-state scalp electroencephalography (EEG) is a promising method for predicting patient outcomes of antidepressant treatments. Machine-learning-based EEG analysis of averaged power features (APF) have predicted antidepressant responders in standalone samples but have not yet significantly impacted clinical care. Here, we applied new approaches for analyzing *transient* spectral event features (SEF) – single, short-lived increases in non-averaged power – in efforts to improve prediction of antidepressant response and yield novel mechanistic biomarkers of readiness to respond. We analyzed resting-state EEG data from the Establishing Moderators and Biosignatures of Antidepressant Response in Clinical Care (EMBARC) trial, fitting linear elastic net models predicting depression score change post-treatment from pre-treatment SEF from frontal channels. We found that a model containing SEF only significantly predicted depression score changes rather than categorical response outcomes, and that its performance was similar to existing published models with APF alone. Additionally, a model including both SEF and APF outperformed both the others, emphasizing the utility of including SEF in models using EEG features for predicting antidepressant response to sertraline. We further investigated the most predictive SEF from the SEF only model, with the objective of revealing channel-level biomarkers to guide mechanistic insight into antidepressant response. We found that pre-treatment frontopolar beta duration was significantly correlated with depression score change, with greater degree of symptom response linked to shorter beta events at baseline. This finding replicates prior work on frontopolar beta duration as a possible biomarker of antidepressant response in rTMS and raises the possibility that beta events may be a cross-modal therapeutic biomarker in antidepressant treatment.

## Introduction

Major depressive disorder (MDD) is a challengingly heterogeneous disorder. Its etiology and course are proposed to differ across the lifespan, its defining symptoms are diverse, and successful treatment is highly individual. To complicate MDD treatment, psychopharmacological therapies, such as the selective serotonin reuptake inhibitor (SSRI) sertraline, typically require several weeks to months of administration before efficacy can be evaluated. Even after an adequate trial period, patients may not experience remission, leading to a subsequent trial of a different antidepressant or the addition of another agent so they can try a combination of medications (Trivedi et al., 2004). Serial treatment trials extend the time patients spend in a depressive episode and increase the resources required for treatment. Justifiably, much work has focused on discovery and development of baseline or early-in-treatment neural biomarkers of therapeutic response in MDD, whether to pharmacological medications (e.g., Leuchter et al., 2009; Trivedi et al., 2016; Warden et al., 2007) or to other treatment modalities (e.g., Pinna et al., 2018; Roelofs et al., 2021; Van Dijk et al., 2022).

Of the methods that could provide such neural biomarkers, scalp electroencephalography (EEG) has been considered a promising option based on cost and accessibility. Over the years, several promising spectral EEG biomarkers have been proposed to predict MDD disease state and its treatment, such as alpha asymmetry, alpha peak frequency and resonance, and theta power and cordance (Arns et al., 2015;

Corlier et al., 2019; Leuchter et al., 2002; Roelofs et al., 2021; Voetterl et al., 2023).These signatures are often derived on the all-trial or all-timepoints average, for event-related and resting-state data respectively. These or a combination of averaged power and connectivity (e.g., power envelope correlation) features have been used from datasets such as the Canadian Biomarker Integration Network in Depression (CAN-BIND; Lam et al., 2016) and Establishing Moderators and Biosignatures of Antidepressant Response in Clinical Care (EMBARC; Trivedi et al., 2016) to build models predictive of binary response (i.e., improvement of at least 50% in depression symptom scores following treatment) or continuous depression score change. In the case of binary classification, the published accuracy of models that successfully predict treatment response with these features ranges from around 64% (CAN-BIND and EMBARC; Schwartzmann et al., 2023) to 86% (from Cohen et al., 2023). Models predicting continuous depression score change with the same features in deep learning models have produced Pearson correlations between predicted and actual change in depression scores in the range of .31 (Jiao et al., 2025) to .61 (Rajpurkar et al., 2020; Wu et al., 2020). However, this degree of accuracy is not sufficient to enhance MDD patient care. To our knowledge, no predictive models have yet led to changes in clinical decision-making, suggesting further improvements are needed. One promising avenue is to add non-averaged EEG features to the predictive models (e.g., Levitt et al., 2020). Many recent studies have shown that in non-averaged EEG data, brain oscillations often emerge as *transient* high-powered bursts or events lasting only up to a few hundred milliseconds. Differences in spectral averages across conditions can arise from a variety of differences in these *transient* Spectral Event Features (SEF), such as rate, amplitude, duration or frequency-span (Shin et al., 2017). Single-trial spectral event characteristics in the 15-29Hz beta frequency band have been shown to predict MDD treatment response in several cases. Baseline frontopolar beta event duration predicts change in scores on the Inventory of Depressive Symptomatology-Self Report (IDS-SR) scale after 5Hz repetitive transcranial magnetic stimulation treatment (rTMS) in MDD patients with comorbid post-traumatic stress disorder (PTSD; Morris et al., 2021). Additionally, baseline beta event rate at multiple frontal and central channels is predictive of improvement in executive function scores after 10Hz rTMS treatment for MDD (Kavanaugh et al., 2023). However, to date, no predictive models of pharmacological antidepressant response have included transient SEF. It is unknown if SEF predict response to antidepressant medications like SSRIs or whether they contain predictive information above and beyond that contained in average spectral characteristics.

In this investigation, we 1) assessed whether pre-treatment SEF can be used to construct a machine-learning model predictive of MDD treatment response to sertraline, and 2) assessed whether models built with pre-treatment SEF outperform those built with commonly used Averaged Power Features (APF). We conducted a secondary analysis of EEG data from Phase 1 of the EMBARC clinical trial data, in which MDD participants received sertraline or placebo for eight weeks (Trivedi et al., 2016). We hypothesized that SEF could be used to successfully predict MDD treatment response to sertraline and that the addition of SEF to predictive models would improve predictability above APF models alone. We also predicted that SEF from our predictive model would correspond to prediction of treatment response at the channel level outside of the model, thus uncovering predictive EEG features in non-averaged data that may be used to infer neuronal and circuit bases of the signals in future investigations.

## Methods

### Participants and Data

We analyzed the EMBARC trial resting-state EEG, symptom scales, and demographic data from the open NIMH Data Archive with a Data Use Agreement. The multi-site clinical trial design is discussed in detail in Trivedi et al. (2016). Briefly, during Phase 1, adults with primary MDD were blindly randomized to receive an eight-week course of placebo or sertraline. Resting-state EEG was collected prior to randomization and during Week 1 of treatment. Assessment of MDD severity was done with the 17-item Hamilton Depression Rating Scale (HAM-D) prior to randomization, weekly, and after the final week of treatment; responder status was defined by ≥ 50% decrease in HAM-D score from baseline to endpoint and remission was defined by endpoint HAM-D score of 7 or less. Resting state EEG was also collected from a small number of healthy control (HC) participants at one time point. We restricted our analyses to participants who had pre-treatment resting-state EEG recordings, post-treatment HAM-D scores, and who had EEG data collected with a 10-20 mapping system (i.e., excluding McLean Hospital site participants with geodesic net caps). This yielded 176 MDD participants and 27 HCs. HC data were used in analyses described below to compare predictive channel-level SEF between depressed and healthy trial participants.

The final analyzed sample included 176 MDD patients (123 female, mean age 38.70) and 27 HCs (17 female, mean age 37.30; age-matched to a subsample of MDD patients for patient-control comparisons, see below). Of the 82 MDD patients assigned to receive sertraline in EMBARC’s Phase I, 43 were sertraline responders (31 female, mean age 40.51) and 39 were sertraline non-responders (29 female, mean age 38.18). Among those assigned to sertraline, 37 were considered in remission (27 female, mean age 40.95) post-treatment, and 45 were not (33 female, mean age 38.13). Of the 94 patients assigned to receive placebo, 58 were responders (40 female, mean age 40.81) and 36 were non-responders (23 female, mean age 33.69).

### Treatment outcomes

The primary measure of treatment outcome used as the predicted value for model training and testing was percentage change in summed 17-item HAM-D score from pre-treatment randomization to post-treatment Week 8. Group comparisons of response and remission rates were run in post-hoc analyses. In those analyses (and in binary classification models described below and in the **Supplement**), response was defined as greater than or equal to 50% decrease in summed HAM-D and non-response as less than 50% decrease.

### EEG analyses

*Preprocessing.* Data were imported into EEGLAB (Delorme & Makeig, 2004) using the appropriate plug-ins for each file format, then highpass-filtered at 0.5Hz and lowpass-filtered at 50Hz. Noisy channels were automatically detected and removed without interpolation based on channel correlation and broken time. Noisy 1s epochs were detected and removed based on outlier statistics and kurtosis. Cleaned data were re-referenced to the common average and subjected to independent components analysis (ICA) with extension to sub-gaussian sources (Bell & Sejnowski, 1995). ICs identified as eye artifacts were removed. Finally, the data from all standard 10-20 frontal electrodes (i.e., Fp1, Fpz, Fp2, Fz, F4, F3, F7, and F8) were epoched into 5s segments before spectral event extraction. These channels were chosen based on prior literature which found predictive biomarkers for 5Hz rTMS treatment in frontal channels (Morris et al., 2021) and to maximize opportunity to validate and replicate these results in other datasets.

*SEF extraction.* SEF were extracted with the SpectralEvents toolbox (https://github.com/jonescompneurolab/SpectralEvents; see **Table 1**). Our event detection method followed Shin et al. (2017). Events were detected by finding maxima in the TFR domain, then selecting suprathreshold (above 6 x factor-of-median) peaks in specific bands (i.e., findMethod = 1). As in prior studies (Shin et al., 2017; Morris et al., 2023), we chose this cutoff to best account for events that relate to the total mean resting-state EEG power over the course of the recording. We calculated in each subject and frequency band the correlation between the power in the area above cutoff (that is, frequencies and time points that would be counted as transient events) in the frequency x time spectrogram and the mean power in the frequency band across all time points for cutoffs of 2, 4, and 6 x median. As shown in **Supplementary Figure 1**, the correlation of event area with mean power was highest near the 6 x median power cutoff in all bands. We note we did not test values above 6 x median due to the very low number of spectral events extracted in some bands at this cutoff level. The find method used in this investigation is the same as in prior work studying SEF therapeutic prediction (Morris et al., 2022) and allows for multiple events to occur in a suprathreshold region. Features extracted included the rate, duration, frequency span, and power of events in the low frequency “delta/theta” band (2-5Hz), alpha band (6-14Hz), and beta band (15-29Hz). These bands were defined based on the visible definition of the closed-eyes alpha power peak and surrounding frequencies in the all-subjects averaged power spectral density (see **Figure 1A**). Each feature was averaged within the band and electrode (for each electrode described in the previous paragraph), across all extracted events for a single subject. This was the same procedure used in Morris et al. (2021) . SEF were then normalized as described below prior to predictive model fitting.

**Figure 1.**
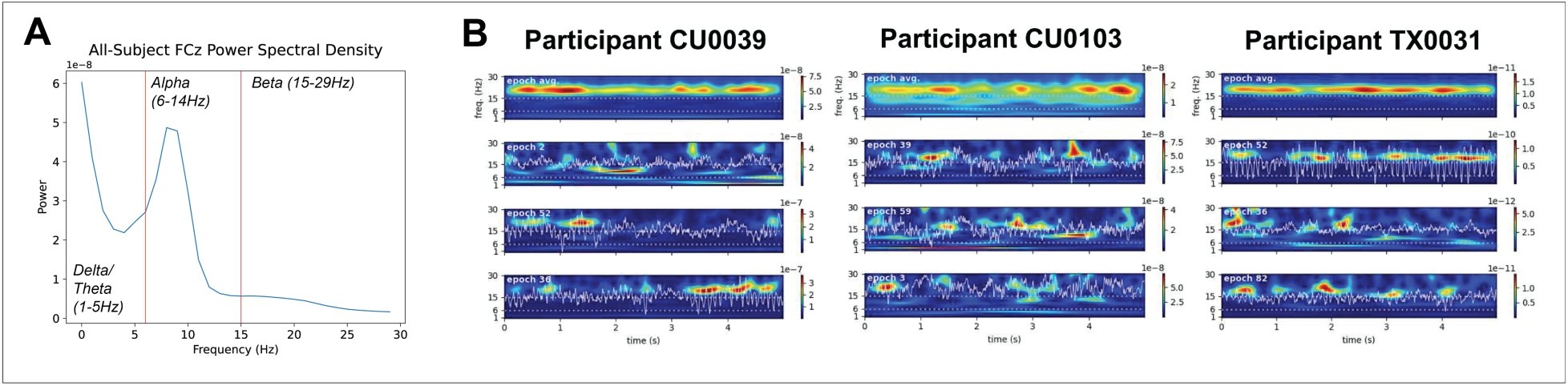
A) Averaged power spectral density of the entire sample and cutoffs for delta/theta, alpha, and beta band definitions. B) Averaged power (top) across all 5 second epochs in the resting state EEG recordings of three example participants, and three example epochs containing event-like transient spectral activity (bottom three).

**Table 1.** Time-frequency EEG signature features extracted for use in predictive models. Each feature was extracted at each of the frontal 10-20 channels (or in the case of connectivity, for each possible pair) in the delta/theta, alpha, and beta bands.

| *Spectral Event Features* | *Averaged Power Features* |
| --- | --- |
| Factor-of-median-normalized power | Absolute power (overall and band-wise) |
| Rate per 5s | Relative band-wise power |
| Full-width half-max duration | Cordance |
| Frequency span | Power envelope pairwise connectivity (PEC) |

*APF extraction.* We extracted the APF (see **Table 1**) most commonly used in machine learning models of treatment response (Jiao et al., 2025; Schwartzmann et al., 2023; Trivedi et al., 2018; Wu et al., 2020) for the purpose of comparing predictability of these features alone against combined models including transient SEF. These features were absolute power across all bands and within delta/theta, alpha, and beta bands (defined as above), relative power in the delta/theta, alpha, and beta bands, cordance in delta/theta, alpha, and beta bands, and power-envelope pairwise connectivity (PEC) across the analyzed frontal 10-20 electrodes in delta/theta, alpha, and beta bands. Relative power was calculated by dividing band-wise averaged power by all-band absolute power. Cordance was defined as the average of z-scored relative and absolute power within each band. PEC was calculated for each pair of channels by deriving the average of correlation coefficients of the two-way orthogonalized amplitude envelopes.

### Elastic net linear prediction models with SEF

Our first objective was to assess whether SEF can be used to predict MDD treatment response, quantified as %change in HAM-D score following sertraline (N=82 active drug group only). To accomplish this, we used linear predictive models. Elastic net (EN) models predicting %change in HAM-D were fit and evaluated using *scikit-learn* (Pedregosa et al., 2011). Features included in the model for each participant were the values of 4 pre-treatment SEF (amplitude, duration, frequency span, and rate, see **Table 1**) in 3 bands (delta/theta, alpha, beta) at 8 frontal channels. Additionally, we included patients’ pre-treatment HAM-D scores to account for baseline depression severity. Additionally, we included patients’ pre-treatment HAM-D scores to account for a potential relationship between pre-treatment symptoms and treatment outcomes (i.e. post-treatment HAM-D scores). Additional sociodemographic variables such as age were not included as features in our models.

Prior to model fitting, feature data was normalized by converting to z-scores, and missing values were interpolated with K-nearest neighbors interpolation based on the 2 nearest-neighbors. Stratified 3-fold cross validation was used to assess model performance. Participants were stratified to ensure the ratio of responders and non-responders in all data subsets approximately matched the ratio present in the full dataset. Participants were randomly split into a training set and a held-out test set (2/3 training and 1/3 testing). Three-fold cross-validation was applied to the training set in combination with a grid search to identify the best model hyperparameters (regularization alpha, L1/L2 ratio, and inclusion of a y intercept). The set of hyperparameters with minimal validation loss (negative root mean squared error) was used for the final model which was evaluated on the held-out test set. The model’s performance in both phases was also tested using the *permutation_test_score* function (which implements a one-sided test under the logic that meaningful prediction should have higher accuracy values than chance) with 5000 permutations. During these permutations, models are re-fit to label-shuffled data to generate a null distribution from which a *p*-value can be calculated to evaluate model performance against chance. In the training set, permutations were performed and averaged over 3 folds (i.e., the same folds used for parameter selection). Permutation testing was performed once on the held-out testing set. This procedure was also repeated on the placebo subgroup (N = 94) to assess whether a model with new parameters could predict placebo response from frontal SEF to test the hypothesis that our EN model with SEF would predict %change in HAM-D only in the active drug group.

### Predictive models with APF

To test whether models including transient SEF outperform binary classification and linear predictive models constructed only with commonly used APF (see **Supplementary Table 1**), we tested the performance of several model types constructed with either 1) APF alone or 2) both averaged power and transient SEF combined (APF+SEF). Classification models tested included K-Nearest Neighbors (KNN) clustering, support vector machines (SVM), and random forests (RFs). These algorithms were chosen based on their prior use with the EMBARC dataset or predicting response in similar patient datasets. For linear prediction, EN was used. Feature normalization, training/validation/testing data subset division, and hyperparameter selection was performed the same as described above for the EN model constructed with transient Spectral Events Features alone. To provide a comparison to non-linear predictive models of %change HAM-D, we also include in **Supplementary Table 2** information about tests with Support Vector Regression, which did not yield significantly predictive models with the non-linear radial basis kernel. To further validate our choice of spectral event detection threshold, we also tested EN models with SEF extracted at different factor-of-median thresholds (see Supplementary Table 3). Only the models using SEF extracted at 6 x median yielded significantly predictive performance.

### Subsequent tests of predictive features

A secondary objective was to identify which of the most predictive channel-level model SEF found in the EN linear prediction model correlated with treatment response outside of the model, with the motivation of identifying localized biomarkers to drive mechanistic understanding of the therapeutic response (see Discussion). To accomplish this, we conducted subsequent analyses on the five most predictive features in our EN model in the sertraline group (N = 82). Feature importance was calculated using the *permutation_importance* function in scikit-learn. Of the resulting SEF in the top five predictors (the fifth was pre-treatment HAM-D score), we examined which were predictive of treatment response outside of the model by calculating the correlation coefficient of the baseline event characteristic values with HAM-D %change, correcting for multiple comparisons across the four features to *p*<.05. For the resulting feature that was significant at the channel level (Fp1 beta duration; see Results), we compared the average beta duration between responders and non-responders, remitters and non-remitters, and MDD patients and HCs with *t*-tests. As there were fewer HCs than MDD patients, a subsampled group of MDD patients matched in age to HCs was used for comparison.

## Results

### Spectral Events are present in the delta/theta, alpha, and beta bands in pre-treatment resting-state data

We first confirmed that event-like spectral activity exists in the non-averaged resting state spectral data in the EMBARC sertraline-treated subsample (N = 82). Examination of time-frequency spectrograms from 1-30Hz averaged across 5 second epochs (71 total epochs on average across participants) indicated that there were peaks in average resting-state power between 2-5 (delta/theta band), 6-14 (alpha band), and 15-29Hz (beta band; i.e., **Fig. 1A** and **Fig. 1B** top panels). Closer examination of individual single epoch spectrograms normalized to median beta power confirmed that, in non-averaged data, high power activity emerged as transient spectral events in each of these bands, laying the foundation for our subsequent analyses. (See **Fig. 1B** bottom panels for examples from three individuals.) The ranges of resulting SEF across the sample are shown in **Supplementary Figure 3**.

### An EN model predicts change in depression symptoms after treatment from baseline SEF only

To assess whether SEF could be used to predict MDD treatment response to sertraline, we fit a linear EN model including pre-treatment frontal SEF (i.e., amplitude, duration, frequency span, and rate) in all frequency bands to post-treatment HAM-D scores from MDD patients who received the active drug. The EN model significantly predicted pre-to-post treatment %change in HAM-D response (i.e., improvement in depression symptoms) above chance (Training/validation negative RMSE: -37.34, *p* = .005; testing negative RMSE: -32.03, *p* = .03; model hyperparameters from grid search: alpha = 3.0, fit_intercept = False, l1_ratio = 0.5; see **Fig. 2A and Table 2**).

**Figure 2.**
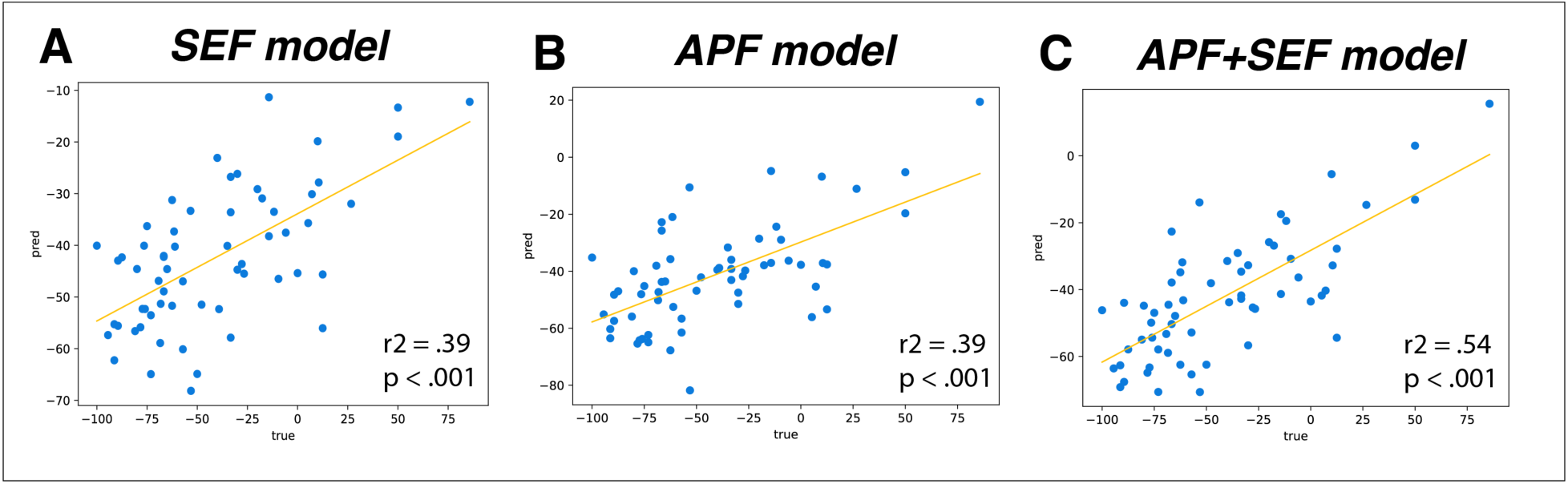
Correlations between trained EN model-predicted versus actual HAM-D %change.

**Table 2.** Parameters and performance of linear prediction models. SERT = sertraline group, PLA = placebo group. Metrics from previous linear models in the literature included are, 1: Wu et al., 2020; 2: Jiao et al., 2025; 3: Rajpurkar et al., 2020. For models tested in the current investigation, p values below .05 indicate the model performed above chance compared to label-shuffled values.

| *Features included* | *Model type* | *Predicted variable* | *NRMSE: Training/Testing* | *P value: Training/Testing* | *R<sup>2</sup> of pred/true HAM-D change in trained model* |
| --- | --- | --- | --- | --- | --- |
| APF only<sup>1</sup> | SELSER | HAM-D change | - | - | .60 |
| APF only + fMRI<sup>2</sup> | DL, graph | HAM-D change | - | - | .31 |
| APF only<sup>3</sup> | Decision trees | HAM-D individual item change | - | - | Up to .55 |
| SEF only, SERT | Elastic net | HAM-D change | -37.34/-32.03 | .005/.03 | .39 |
| SEF only, PLA | Elastic net | HAM-D change | -39.76/-49.48 | .57/.46 |  |
| APF, SERT | Elastic net | HAM-D change | -40.37/-37.34 | .11/.22 | .39 |
| APF+SEF, SERT | Elastic net | HAM-D change | -39.63/-32.96 | .03/.047 | .54 |

To test whether this model was selectively predictive of %change in symptoms after sertraline and not placebo, we used the model fit in the sertraline group to attempt to predict HAM-D %change after 8 weeks of placebo (N = 94) with baseline frontal SEF. The model’s predictions of %change were not significantly above chance for the placebo subgroup (negative RMSE: -43.15, *p* = .68). Furthermore, we could not produce above-chance predictions in the placebo group from baseline SEF, even when constructing a new model from scratch with the same methods used in the active drug group (Training/validation negative RMSE: -39.76, *p* = .57; testing negative RMSE: -49.48, *p* = .46; parameters: alpha = 3.0, fit_intercept = True, l1_ratio = 0.1), suggesting that frontal SEF were selectively predictive of sertraline-associated depression symptom improvement, and not generalizable to the placebo effect.

We compared the EN model’s performance with SEF only to previously published linear models predicting SSRI (sertraline in EMBARC; sertraline in T-RAD, Trivedi et al., 2020; sertraline and escitalopram in iSPOT-D, Williams et al., 2011) treatment response using *only APF* in this or similar data sets (Jiao et al., 2025, Rajpurkar et al., 2020; Wu et al., 2020). To assess whether this model’s performance was comparable to these prior published models, we computed *r^2^* between predicted and true HAM-D percentage changes and compared to reported *r^2^* values in the previous publications. Our trained SEF EN model had a *r^2^* of 0.39 (Pearson’s *r* = .62, *p* < .0001; see **Fig. 2A**). This *r^2^* fell squarely within the range of previously published values from similar models (0.31 < *r^2^* < 0.6, see **Table 2**).

### The inclusion of SEF improves performance of treatment response prediction models compared to models constructed with only APF

Next, we tested if an EN model constructed with APF only, or with both APF and SEF, had improved performance compared to the EN model with SEF only. We found that EN models with APF only could not predict HAM-D %change above chance (model hyperparameters: alpha = 3.0, fit_intercept = True, l1_ratio = 0.1, training *p* = .11, testing *p* = .22, **Table 2**), though the model did show a correlation between predicted and true HAM-D %change in the *training* set (*r^2^* value of .39, **Fig. 2B**). However, an EN model with APF+SEF was significantly predictive of HAM-D %change (alpha = 3.0, fit_intercept = False, l1_ratio = 0.5, training *p* = .03, testing *p* = .047). This model showed an even stronger correlation between predicted and true scores in the training set, with an *r^2^* value of .54 (see **Table 2** and **Fig. 2C**), which is also within the range of prior published models.

We also compared the performance of the linear EN models (**Fig. 2 and Table 2**) to the performance of standard binary treatment response classification models, including those with non-linear feature combination (namely KNN, SVM and RF, see Methods). We first examined if these binary classification models could predict categorical treatment response using baseline SEF only and found that they could not. These models were also not predictive when using Average Power Features alone or both Average Power and SEF (see **Supplementary Table 1**).

### Frontopolar beta event duration predicts improvement in depression scores outside of the SEF EN model

Given the success of the SEF only EN model in predicting HAM-D %change from pre-treatment spectral events alone (**Table 2**), we further examined which SEF were most predictive of post-treatment improvement in depression scores. As discussed in the Introduction, EEG oscillations emerge as short-lived, high-powered bursts or events (such as SEF), whose dynamics can cause changes in APF across groups for a number of reasons (e.g., differences in rate, amplitude, duration or frequency-span). Therefore, quantifying SEF directly provided a finer-grained description of EEG biomarkers of treatment response that cannot be provided by predictive APF features. This is important because SEF and can be more readily linked to the underlying neural generators (see Discussion). We extracted the five most predictive model features from our SEF only EN model and tested whether any of the spectral event characteristics were predictive of depression symptom improvement on their own, outside of the EN model. The top predictive features included F4 theta power and duration, Fz beta duration, Fp1 beta duration, and pre-treatment HAM-D, which all had an absolute perm_importance weight greater than 1.5 (**Fig. 3A**). Of the top SEF predictors, only Fp1 pre-treatment beta duration significantly predicted symptom improvement after correction, such that shorter durations corresponded with greater symptom %change (decrease) in HAM-D; **Fig. 3B**, *p*<0.05).

**Figure 3.**
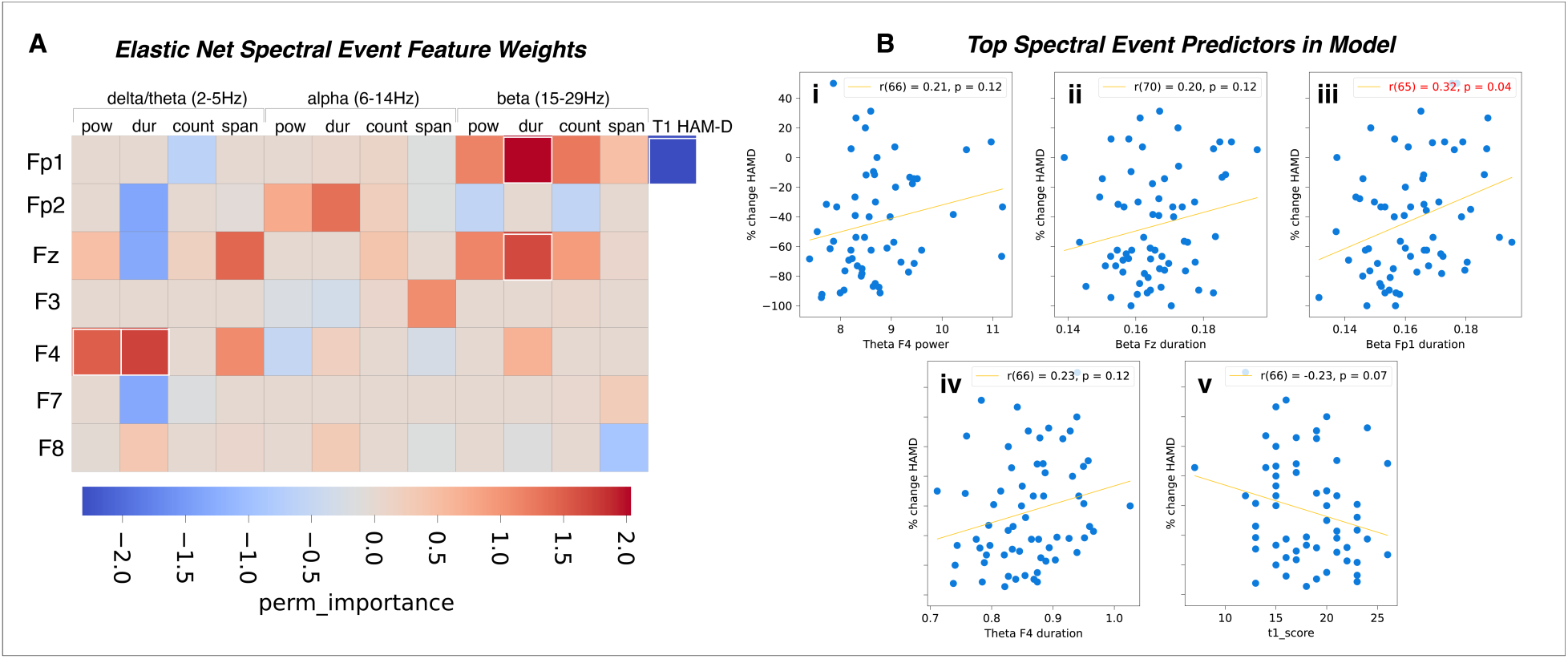
Frontal transient SEF predictive of symptom improvement following sertraline. A) Model weights of the SEF included in the trained EN model. The features included in the top five most predictive are highlighted in white boxes. B) Correlation of the most predictive features’ channel data with HAM-D %change with correction for multiple comparisons.

### Frontopolar pre-treatment beta event duration distinguishes sertraline responders from non-responders, but not HC from MDD

Given the significant correlation between pre-treatment Fp1 beta event duration and symptom improvement, we further tested if Fp1 beta duration significantly differed between 1) sertraline responders and nonresponders, and 2) patients with MDD and HCs. Pre-treatment Fp1 beta duration was significantly shorter in responders and remitters, compared to non-responders and non-remitters, respectively (*t* = -1.71, *p* = .046; *t* = - 1.78, *p* = 0.04; see **Fig. 4A**). However, pre-treatment Fp1 beta duration did not correlate with pre-treatment HAM-D score (*r* = .06, *p* = .63; See **Fig. 4B**), suggesting that this was not simply an artifact of MDD severity at baseline, which prior studies suggest may moderate treatment response (Driessen et al., 2010).

**Figure 4.**
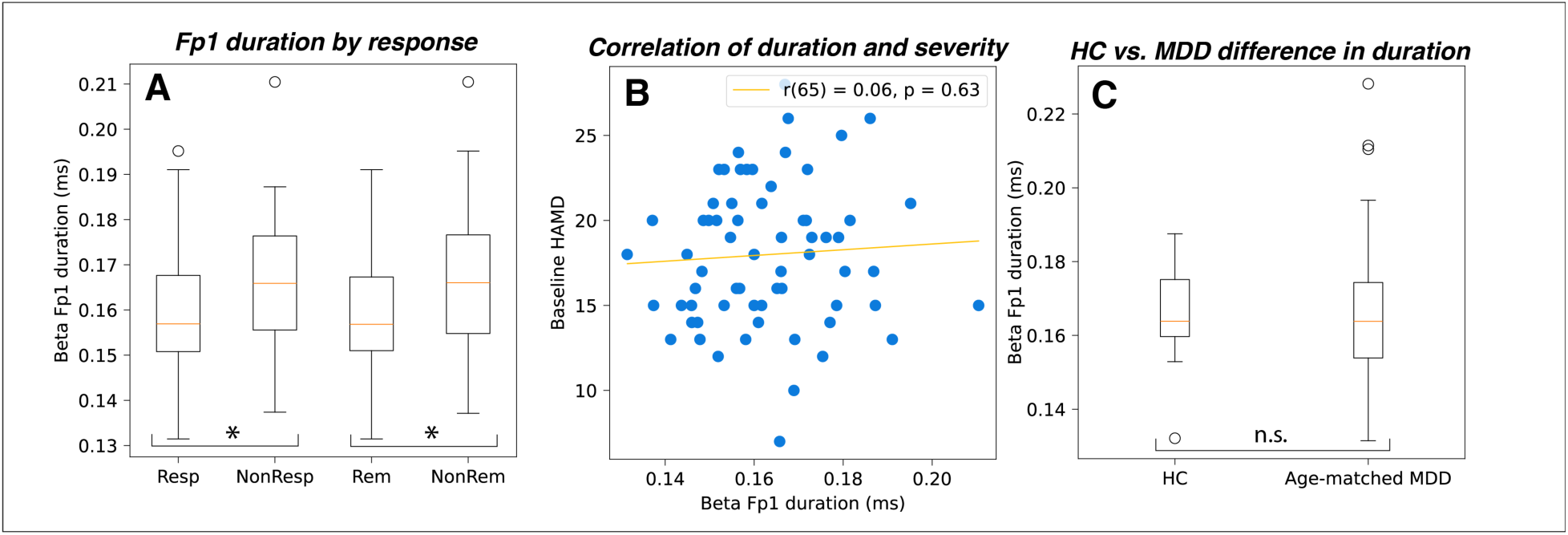
A) Fp1 beta duration is shorter in responders than non-responders and shorter in remitters than non-remitters. B) Fp1 beta duration does not correlate with baseline symptom severity. C) Fp1 beta duration does not differ between MDD and HC groups.

The EMBARC dataset only contains 27 HC. Given the small sample size, to explore whether pre-treatment Fp1 beta duration is also a marker of MDD disease state, we compared HC to a randomly selected, age-matched subset of the MDD patients (27 subjects in each group). We found pre-treatment Fp1 beta duration did not differ between MDD and HC (*t* = 0.24, *p* = .81, see **Fig. 4C**).

## Discussion

### Frontal SEF predicted treatment response in the EMBARC dataset and outperformed models with APF alone

In this secondary analysis of the EMBARC trial resting-state EEG data, our objectives were to investigate whether SEF can be used to predict MDD patients’ response to sertraline treatment above and beyond classic approaches with APF, and to identify which SEF provided the most predictive biomarkers at the channel-level. We found that a machine learning model containing only SEF from the delta/theta, alpha, and beta bands at frontal channels in pre-treatment resting-state EEG successfully predicted continuous depression score %change following sertraline, but not placebo treatment. This model performed equivalently to one built using APF alone, and its prediction accuracy was comparable to the range of predictive values from previously reported significant machine learning models (**Table 2**). A model with both APF *and* SEF outperformed those with one or the other, highlighting the utility of transient SEF in machine learning-based prediction of treatment response. Among the most predictive features of our spectral events-based model was frontopolar beta event duration, which was the only feature that correlated with treatment response at the channel-level. Though pre-treatment beta event durations did not differ between depressed patients and HC (albeit in a small sub-sample) and did not change after one week of sertraline treatment, they were significantly shorter in eventual responders and remitters than non-responders and non-remitters, yielding a novel biomarker of antidepressant response.

### SEF boosted predictive power in treatment-predicting models

Much work has already been done to identify APF that are predictive of treatment response in MDD in multiple treatment modalities (see review by Olbrich & Arns, 2013), such as frontal theta power (Arns et al., 2015), theta cordance (Leuchter et al., 2002), individual alpha peak frequency (Corlier et al., 2019; Voetterl et al., 2023), and proximity of individual peak alpha frequency to rTMS stimulation frequency (Roelofs et al., 2021). Models constructed with APF have also successfully identified cognitive patient phenotypes within the EMBARC dataset, wherein changes in band-specific source-localized power related to specific deficits in task performance (e.g., blunted reinforcement learning, cognitive control deficits, etc., Webb et al., 2016). All these prior studies have relied on averaging resting-state power features. A growing body of work has shown that averaged resting-state power is driven by aspects of transient SEF in the non-averaged data, including the rates, amplitude and duration of the spectral events (e.g., Kavanaugh et al., 2023; Levitt et al., 2020; Morris et al., 2021; Shin et al., 2017; see **Fig. 1, Supp. Fig. 3**). While including both SEF and APF in a predictive machine learning model may seem redundant, our results show that inclusion of both leads to better predictions of improvement in depression severity following sertraline treatment than models with either SEF or APF alone. As such, it is possible that the inclusion of transient SEF in preexisting published models of treatment response (e.g., see list in **Table 2**) may boost their predictive power.

### Beta event duration may be a biomarker for sertraline-or other treatment-responsive MDD

Though it is tempting at first glance to speculate that sertraline may work by “re-normalizing” beta events to a “healthy” length, our subsample analysis showed Fp1 beta event duration was not significantly longer in HCs compared to an age-matched subsample of EMBARC MDD patients. Furthermore, Fp1 beta event duration did not change significantly from pre-treatment to post-Week 1 (see **Supplementary Figure 4**), though this may be due to the fact that the therapeutic effects of sertraline on depressive symptoms had not yet occurred. Taken together, this evidence suggests that Fp1 beta duration may be considered a biomarker of the specific type of neural pathology that will respond to sertraline therapy.

Importantly, frontopolar beta duration has been shown to be a potential biomarker of readiness to respond to another MDD treatment modality – rTMS (Morris et al. 2021). Fp2 and Fpz beta duration were similarly predictive of %change in depression severity scores after a course of 5Hz rTMS in patients with comorbid MDD and PTSD. Additionally, the same direction of correlation between beta duration and symptoms was found in that study, that is, with shorter beta events predicting greater improvement in depression symptoms. With a post-treatment EEG recording, they were able to demonstrate that the change in beta durations at Fp1, Fp2, and Fpz were significantly correlated with depression score change, with beta event duration increasing with greater symptom reduction. Our replication of this result in sertraline treatment (albeit restricted to the baseline timepoint) suggests that frontopolar beta event duration may be a reproducible marker of underlying “readiness to respond” factors that generalize across treatment modalities in MDD. One possibility is that beta event duration is indicative of baseline plasticity that improves treatment response. Both D1 activation (Enomoto et al., 2015) and NMDA partial agonists (Brown et al., 1999) have been shown to increase the long-term potentiation effects of TMS. Baseline beta event duration may reflect natural variation in baseline plasticity in these receptors among MDD patients, which could guide treatment choice or perhaps be augmented, as is being attempted in accelerated rTMS treatment courses that include administration of NMDA partial agonists or stimulants to boost plasticity (Vaughn et al., 2024).

### Leveraging SEF to yield mechanistic insights into antidepressant treatment response

Another notable aspect of our beta duration finding is that the neural mechanisms of beta event generation have been explored. Prior studies combining biophysical neural modeling, human MEG imaging, and invasive animal recordings (Bonaiuto et al., 2021; Law et al., 2022; Sherman et al., 2016) lend insights into additional mechanistic factors that could help explain why beta event duration is predictive of antidepressant response. These studies employed a biophysical modeling software called Human Neocortical Neurosolver (HNN; Neymotin et al., 2020). HNN’s core is a model of the canonical neocortical column under exogenous drive designed to simulate multiscale mechanisms that lead to macroscale human M/EEG signals, such as transient spectral events. Sherman et al. (2016) used this model alongside MEG and laminar recordings from non-human models to demonstrate that characteristics of layer-specific thalamic input to cortex likely controls beta event generation. This beta-generating thalamic drive has also been proposed to recruit cortical long-lived inhibition (Law et al., 2022). Together, this evidence raises the possibility that thalamocortical signaling and inhibitory tone in the cortex might be related to beta duration and differ between antidepressant responders and non-responders. To better understand whether beta event durations relate to *treatment effects* mediated by thalamocortical signaling or inhibitory tone, future investigations should address whether beta event increases are observed after a full course of sertraline.

### Limitations of the current investigation

There were several limitations inherent to this investigation. First, the dissociation between timepoints at which EEG data were collected (before and one week into treatment) and the time course of SSRI therapeutic effects (typically 6-8 weeks) meant that we were unable to report how SEF relate to treatment mechanism. In the future, this might be addressed through the investigations using faster-acting therapies such as esketamine or accelerated TMS protocols that could reveal EEG biomarker changes more specific to the mechanism of treatment. Second, because the scope of the current study was the identification of SEF biomarkers of treatment response, we did not include sociodemographic variables as features in predictive models. Third, though we performed within-sample held-out testing, it is possible that elastic net models including SEF may not generalize to predict treatment response in other independent samples. Finally, the small sample size in the healthy control group limits the power of our depressed-control comparison. These factors should be considered in future prospective study design and in construction and replication of predictive models including SEF.

In summary, future work should continue to investigate whether beta event duration is indicative of baseline plasticity that impacts readiness to respond to antidepressant treatment. Shifting from using APF to spectral and waveform features of transient spectral events may lead to more predictive models, new clinical biomarkers, and insights into the mechanisms of antidepressant response.

## Data Availability

All statistical and analysis code in the present study are available upon reasonable request to the authors, which can be used to recreate results of the present work, though approval through the NIH Data Archive is necessary to access EMBARC study data.

## Acknowledgements

This work was supported by funding from the NIH (T32 MH126388 to DAW; MH130415 to SRJ; 5R01MH135293 to LLC). The authors would like to thank Dr. Nicholas Tolley for his input on description of the machine learning methods in this article.

## Supplementary Information

### Determination of Factor-of-Median (FOM) threshold cutoff

**Supplementary Figure 1.**
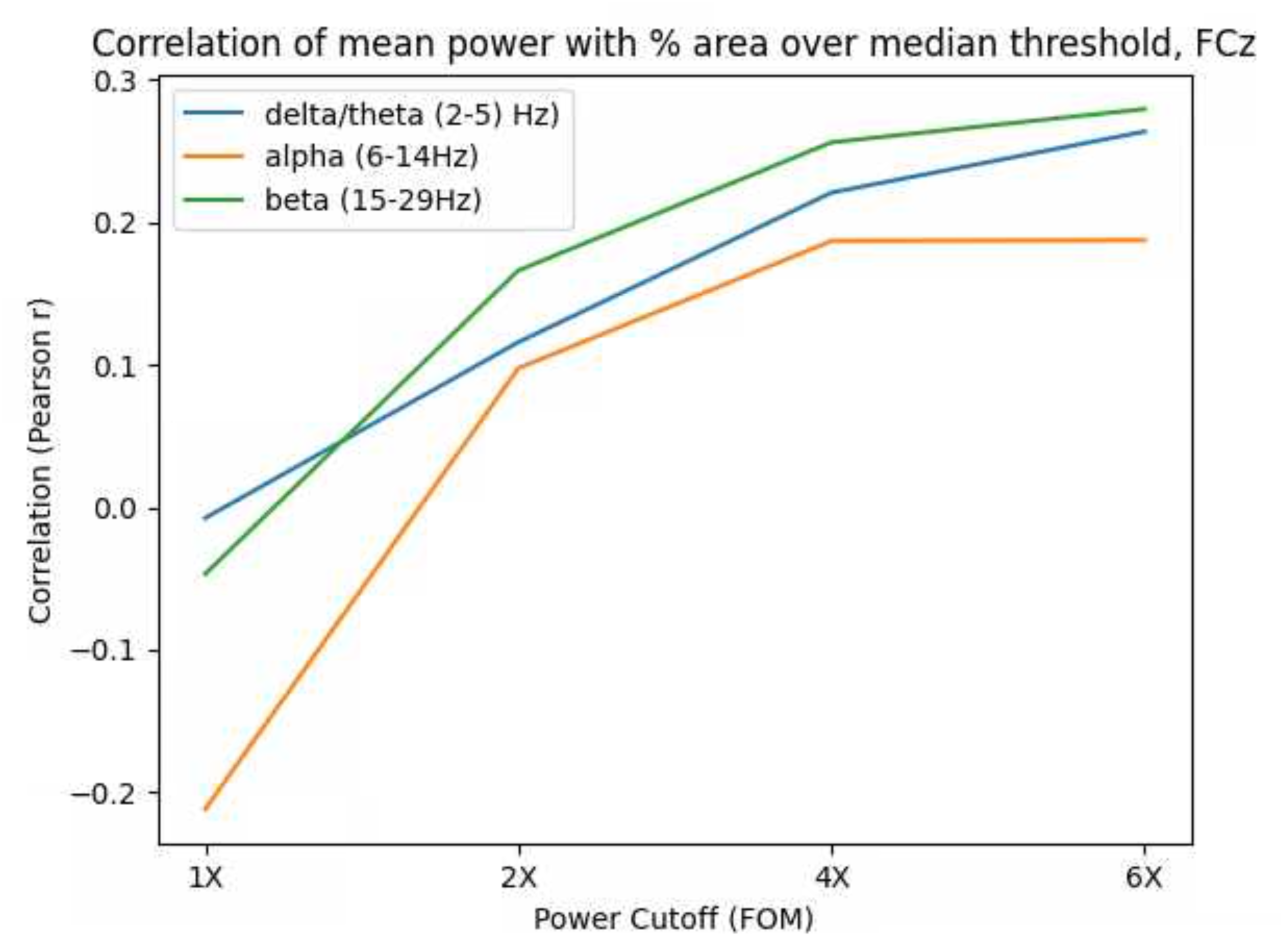
Correlation between the power in the area above cutoff (that is, frequencies and time points that would be counted as transient events) in the frequency x time spectrogram and mean power in the frequency bands of interest. 6 x FOM had the highest correlation with mean power in all frequency bands, validating this choice of cutoff in the current study.

### Model training, validation, and testing

**Supplementary Figure 2.**
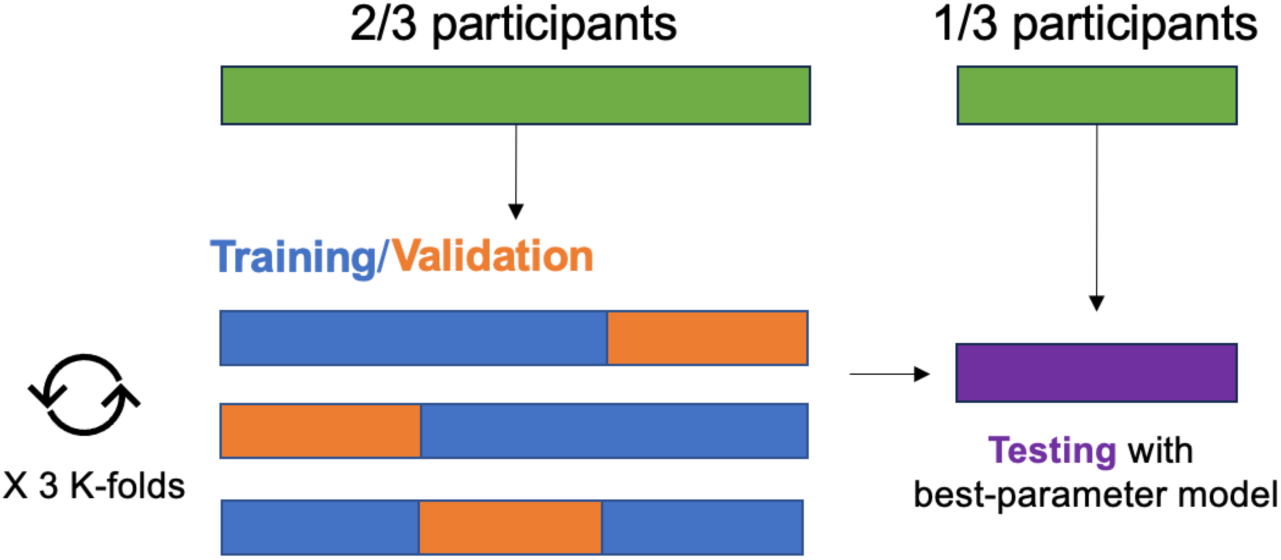
Diagram of model training/validation and testing procedures. The cross-validated grid search and training/validation procedure was performed on 2/3 of participants using 3 K-folds for cross-validation. Testing was performed on the held-out 1/3 of participants using the resulting model with the best hyperparameters.

### Range of SEF in Sertraline Group

**Supplementary Figure 3.**
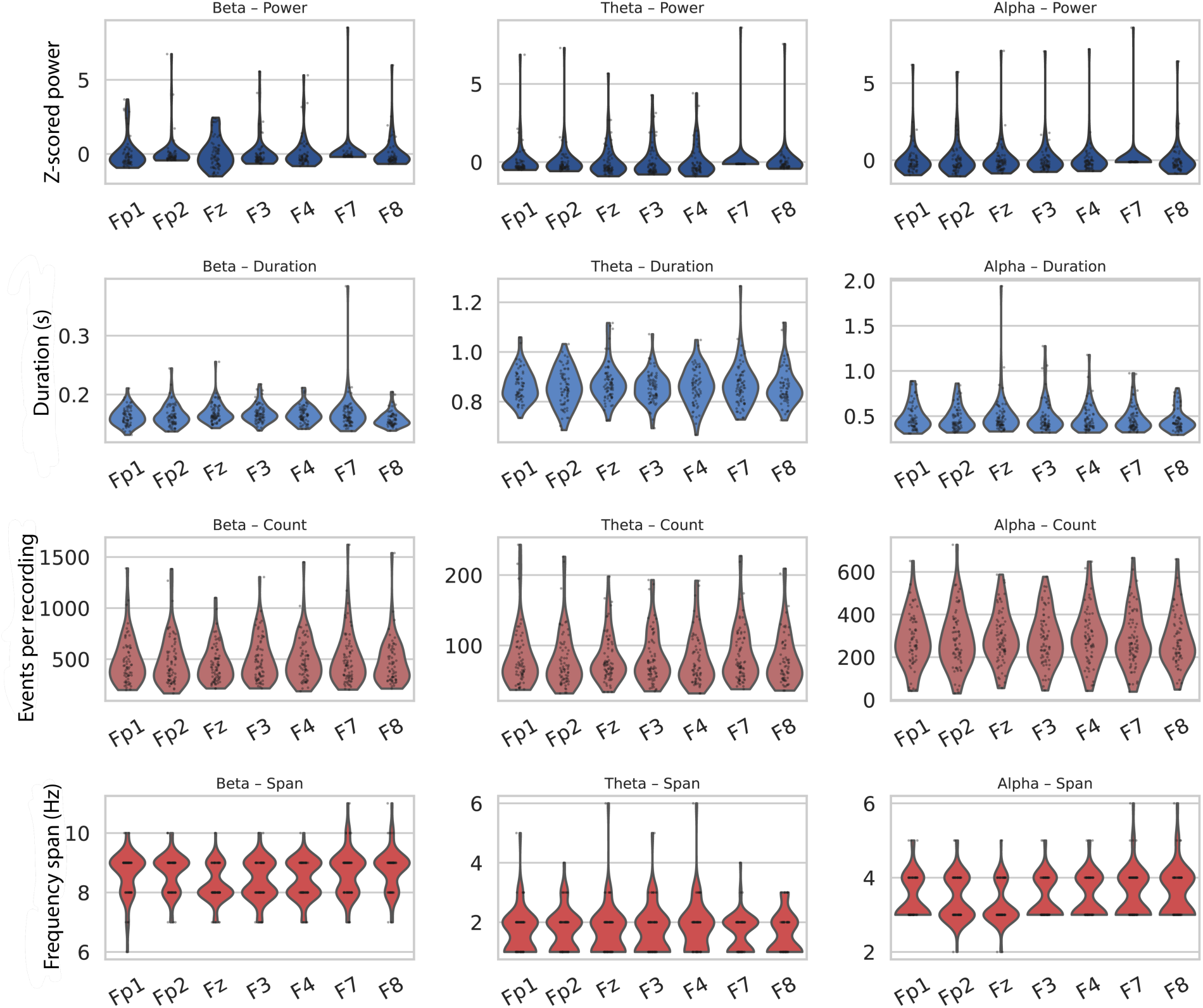
Range of SEF at each channel and band for the subsample (N = 82) who received sertraline treatment.

### Models cannot significantly classify binary sertraline treatment response/nonresponse with SEF only, APF, or SEF+APF

As described in the Methods and Results section, classification models predicting categorical (responder vs. non-responder) treatment outcomes from baseline SEF did not perform significantly above chance. For completeness and to compare performance against models presented in the literature, we also tested the performance of classification models constructed with Average Power Features only and with both SEF and APF. None of the classification models tested performed above chance at predicting sertraline responder status in the EMBARC dataset (see **Supplementary Table 1**).

**Supplementary Table 1.** Accuracy and significance of published models of binary classification models of antidepressant response and classification models tested in the current study. 1: <u>Schwartzmann et al., 2023</u>; 2: <u>Zhdanov et al., 2020</u>; 3: <u>Jaworska et al., 2019</u>.

| *Features included* | *Model type* | *Predicted variable* | *Accuracy (F1):*<br>*Training/Testing* | *P value:*<br>*Training/Testing* |
| --- | --- | --- | --- | --- |
| APF only<sup>1</sup> | KNN | Response (y/n) | .64 | - |
| APF only<sup>2</sup> | SVM | Response (y/n) | .79 | - |
| APF only<sup>3</sup> | RF | Response (y/n) | .88 | - |
| SEF only | SVM | Response (y/n) | .54/.58 | .27/.17 |
| APF | SVM | Response (y/n) | .54/.35 | .29/.70 |
| APF+SEF | SVM | Response (y/n) | .49/.60 | .53/.14 |
| SEF only | KNN | Response (y/n) | .02/.42 | .64/.45 |
| APF | KNN | Response (y/n) | .60/.39 | .09/.78 |
| APF+SEF | KNN | Response (y/n) | .59/.38 | .07/.56 |
| SEF only | RF | Response (y/n) | .93/.94 | .37/.22 |
| APF | RF | Response (y/n) | .52/.61 | .38/.13 |
| APF+SEF | RF | Response (y/n) | .45/.22 | .68/.95 |

### Support Vector Regression does not significantly predict treatment response with the non-linear radial basis kernel with SEF, APF, or both

To assess whether models that implement non-linear combinations of features instead (as opposed to linear combinations in our EN model) 1) significantly predict %change in HAM-D from SEF and/or 2) have performance improved by the inclusion of SEF, we constructed and tested the performance of Support Vector Regression (SVR) models using the radial basis kernel. SVR did not perform above chance at predicting %change HAM-D with APF, or a combination of both SEF and APF. However, SVR with SEF alone yielded a significantly predictive training-set model, though the testing-set model did not quite reach significance. This could indicate potential utility of non-linear combinations of SEF in predicting treatment response.

**Supplementary Table 2.**
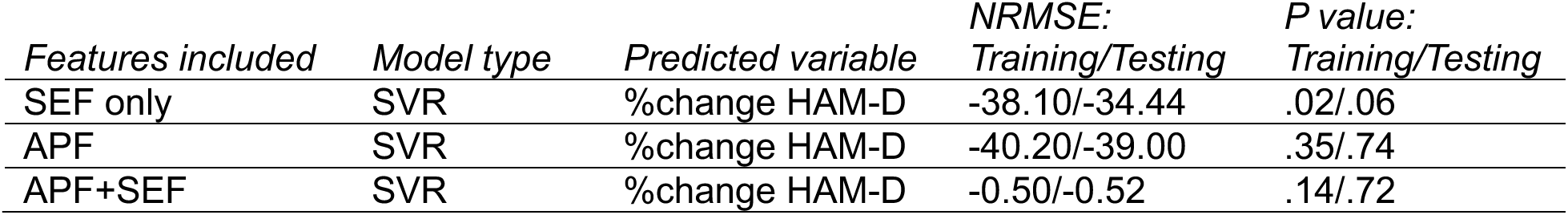
Accuracy and significance of a non-linear model of %change HAM-D (i.e., support vector regression with the radial basis kernel) tested in the current study.

### Sensitivity of model outcomes to factor-of-median SEF detection threshold

To test the effect of chosen factor-of-median detection threshold used to quantify spectral events in the SpectralEvents toolbox, we tested model performance when using SEF derived from events quantified at 4 x and 2 x factor-of-median power. We found that models including SEF quantified at these thresholds did not successfully generalize to predict above-chance in the validation stage.

**Supplementary Table 3.** Accuracy and significance of elastic net models with SEF extracted using different factor-of-median detection thresholds.

| *Features included* | *Model type* | *Predicted variable* | *NRMSE:*<br>*Training/Testing* | *P value:*<br>*Training/Testing* |
| --- | --- | --- | --- | --- |
| SEF only (6xFOM) | EN | %change HAM-D | -37.34/-32.03 | .005/.03 |
| SEF only (4xFOM) | EN | %change HAM-D | -37.17/-35.68 | .004/.13 |
| SEF only (2xFOM) | EN | %change HAM-D | -36.76/-40.77 | .002/.40 |

### Frontopolar beta duration does not differ between pre-treatment baseline and 1 week on sertraline

While baseline Fp1 duration is predictive of post-treatment symptom changes after eight weeks of treatment, we did not see a significant change in beta duration one week into treatment (t = -1.27, p = .22), which is the only other timepoint at which EEG was collected in this study.

**Supplementary Figure 4.**
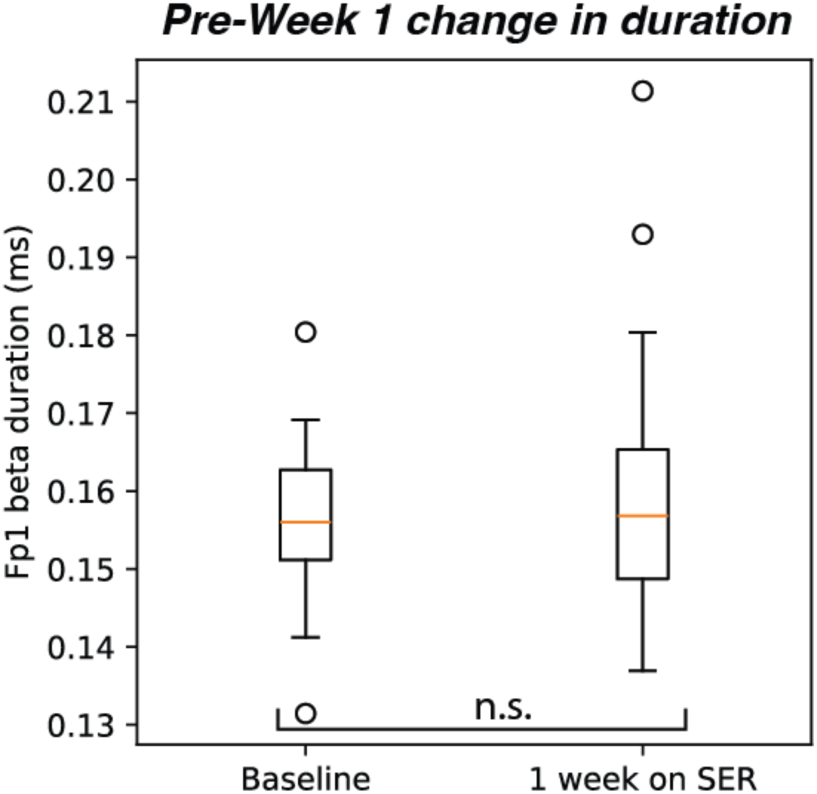
Fp1 beta duration does not significantly change during one week of sertraline treatment. * = p < .05; n.s. = not significant.

## References

Arns, M., Etkin, A., Hegerl, U., Williams, L. M., DeBattista, C., Palmer, D. M., Fitzgerald, P. B., Harris, A., deBeuss, R., & Gordon, E. (2015). Frontal and rostral anterior cingulate (rACC) theta EEG in depression: Implications for treatment outcome? European Neuropsychopharmacology, 25(8), 1190–1200. 10.1016/j.euroneuro.2015.03.007

Bell, A. J., & Sejnowski, T. J. (1995). An Information-Maximization Approach to Blind Separation and Blind Deconvolution. Neural Computation, 7, 31.

Bonaiuto, J. J., Little, S., Neymotin, S. A., Jones, S. R., Barnes, G. R., & Bestmann, S. (2021). Laminar dynamics of high amplitude beta bursts in human motor cortex. NeuroImage, 242, 118479. 10.1016/j.neuroimage.2021.118479

Brown, R. G., Limousin Dowsey, P., Brown, P., Jahanshahi, M., Pollak, P., Benabid, A. L., Rodriguez-Oroz, M. C., Obeso, J., & Rothwell, J. C. (1999). Impact of deep brain stimulation on upper limb akinesia in Parkinson’s disease. Annals of Neurology, 45(4), 473–488. 10.1002/1531-8249(199904)45:4<473::AID-ANA9>3.0.CO;2-V

Cohen, S. E., Zantvoord, J. B., Wezenberg, B. N., Daams, J. G., Bockting, C. L. H., Denys, D., & van Wingen, G. A. (2023). Electroencephalography for predicting antidepressant treatment success: A systematic review and meta-analysis. Journal of Affective Disorders, 321, 201–207. 10.1016/j.jad.2022.10.042

Corlier, J., Carpenter, L. L., Wilson, A. C., Tirrell, E., Gobin, A. P., Kavanaugh, B., & Leuchter, A. F. (2019). The relationship between individual alpha peak frequency and clinical outcome with repetitive Transcranial Magnetic Stimulation (rTMS) treatment of Major Depressive Disorder (MDD). Brain Stimulation, 12(6), 1572–1578. 10.1016/j.brs.2019.07.018

Delorme, A., & Makeig, S. (2004). EEGLAB: An open source toolbox for analysis of single-trial EEG dynamics including independent component analysis. Journal of Neuroscience Methods, 134(1), 9–21. 10.1016/j.jneumeth.2003.10.009

Driessen, E., Cuijpers, P., Hollon, S. D., & Dekker, J. J. M. (2010). Does pretreatment severity moderate the efficacy of psychological treatment of adult outpatient depression? A meta-analysis. Journal of Consulting and Clinical Psychology, 78(5), 668–680. 10.1037/a0020570

Enomoto, H., Terao, Y., Kadowaki, S., Nakamura, K., Moriya, A., Nakatani-Enomoto, S., Kobayashi, S., Yoshihara, A., Hanajima, R., & Ugawa, Y. (2015). Effects of l-Dopa and pramipexole on plasticity induced by QPS in human motor cortex. Journal of Neural Transmission, 122(9), 1253–1261. 10.1007/s00702-015-1374-8

Jaworska, N., de la Salle, S., Ibrahim, M.-H., Blier, P., & Knott, V. (2019). Leveraging Machine Learning Approaches for Predicting Antidepressant Treatment Response Using Electroencephalography (EEG) and Clinical Data. Frontiers in Psychiatry, 9. 10.3389/fpsyt.2018.00768

Jiao, Y., Zhao, K., Wei, X., Carlisle, N. B., Keller, C. J., Oathes, D. J., Fonzo, G. A., & Zhang, Y. (2025). Deep graph learning of multimodal brain networks defines treatment-predictive signatures in major depression. Molecular Psychiatry, 30(9), 3963–3974. 10.1038/s41380-025-02974-6

Kavanaugh, B. C., Fukuda, A. M., Gemelli, Z. T., Thorpe, R., Tirrell, E., Vigne, M., Jones, S. R., & Carpenter, L. L. (2023). Pre-treatment frontal beta events are associated with executive dysfunction improvement after repetitive transcranial magnetic stimulation for depression: A preliminary report. Journal of Psychiatric Research, 168, 71–81. 10.1016/j.jpsychires.2023.10.024

Lam, R. W., Milev, R., Rotzinger, S., Andreazza, A. C., Blier, P., Brenner, C., Daskalakis, Z. J., Dharsee, M., Downar, J., Evans, K. R., Farzan, F., Foster, J. A., Frey, B. N., Geraci, J., Giacobbe, P., Feilotter, H. E., Hall, G. B., Harkness, K. L., Hassel, S., … on behalf of the CAN-BIND Investigator Team. (2016). Discovering biomarkers for antidepressant response: Protocol from the Canadian biomarker integration network in depression (CAN-BIND) and clinical characteristics of the first patient cohort. BMC Psychiatry, 16(1), 105. 10.1186/s12888-016-0785-x

Law, R. G., Pugliese, S., Shin, H., Sliva, D. D., Lee, S., Neymotin, S., Moore, C., & Jones, S. R. (2022). Thalamocortical Mechanisms Regulating the Relationship between Transient Beta Events and Human Tactile Perception. Cerebral Cortex, 32(4), 668–688. 10.1093/cercor/bhab221

Leuchter, A. F., Cook, I. A., Marangell, L. B., Gilmer, W. S., Burgoyne, K. S., Howland, R. H., Trivedi, M. H., Zisook, S., Jain, R., McCracken, J. T., Fava, M., Iosifescu, D., & Greenwald, S. (2009). Comparative effectiveness of biomarkers and clinical indicators for predicting outcomes of SSRI treatment in Major Depressive Disorder: Results of the BRITE-MD study. Psychiatry Research, 169(2), 124–131. 10.1016/j.psychres.2009.06.004

Leuchter, A. F., Cook, I. A., Witte, E. A., Morgan, M., & Abrams, M. (2002). Changes in Brain Function of Depressed Subjects During Treatment With Placebo. American Journal of Psychiatry, 159(1), 122–129. 10.1176/appi.ajp.159.1.122

Levitt, J., Edhi, M. M., Thorpe, R. V., Leung, J. W., Michishita, M., Koyama, S., Yoshikawa, S., Scarfo, K. A., Carayannopoulos, A. G., Gu, W., Srivastava, K. H., Clark, B. A., Esteller, R., Borton, D. A., Jones, S. R., & Saab, C. Y. (2020). Pain phenotypes classified by machine learning using electroencephalography features. NeuroImage, 223, 117256. 10.1016/j.neuroimage.2020.117256

Morris, A., Temereanca, S., Zandvakili, A., Sliva, D., Tyrka, A., Greenberg, B., Carpenter, L., Philip, N., & Jones, S. (2021). Transient neural oscillations reveal new biomarkers for therapeutic transcranial magnetic stimulation for comorbid MDD and PTSD. Brain Stimulation, 14(6), 1643. 10.1016/j.brs.2021.10.176

Neymotin, S. A., Daniels, D. S., Caldwell, B., McDougal, R. A., Carnevale, N. T., Jas, M., Moore, C. I., Hines, M. L., Hämäläinen, M., & Jones, S. R. (2020). Human Neocortical Neurosolver (HNN), a new software tool for interpreting the cellular and network origin of human MEG/EEG data. eLife, 9, e51214. 10.7554/eLife.51214

Olbrich, S., & Arns, M. (2013). EEG biomarkers in major depressive disorder: Discriminative power and prediction of treatment response. International Review of Psychiatry, 25(5), 604–618. 10.3109/09540261.2013.816269

Pedregosa, F., Varoquaux, G., Gramfort, A., Michel, V., Thirion, B., Grisel, O., Blondel, M., Prettenhofer, P., Weiss, R., Dubourg, V., Vanderplas, J., Passos, A., & Cournapeau, D. (2011). Scikit-learn: Machine Learning in Python. MACHINE LEARNING IN PYTHON.

Pinna, M., Manchia, M., Oppo, R., Scano, F., Pillai, G., Loche, A. P., Salis, P., & Minnai, G. P. (2018). Clinical and biological predictors of response to electroconvulsive therapy (ECT): A review. Neuroscience Letters, 669, 32–42. 10.1016/j.neulet.2016.10.047

Rajpurkar, P., Yang, J., Dass, N., Vale, V., Keller, A. S., Irvin, J., Taylor, Z., Basu, S., Ng, A., & Williams, L. M. (2020). Evaluation of a Machine Learning Model Based on Pretreatment Symptoms and Electroencephalographic Features to Predict Outcomes of Antidepressant Treatment in Adults With Depression: A Prespecified Secondary Analysis of a Randomized Clinical Trial. JAMA Network Open, 3(6), e206653. 10.1001/jamanetworkopen.2020.6653

Roelofs, C. L., Krepel, N., Corlier, J., Carpenter, L. L., Fitzgerald, P. B., Daskalakis, Z. J., Tendolkar, I., Wilson, A., Downar, J., Bailey, N. W., Blumberger, D. M., Vila-Rodriguez, F., Leuchter, A. F., & Arns, M. (2021). Individual alpha frequency proximity associated with repetitive transcranial magnetic stimulation outcome: An independent replication study from the ICON-DB consortium. Clinical Neurophysiology, 132(2), 643–649. 10.1016/j.clinph.2020.10.017

Schwartzmann, B., Dhami, P., Uher, R., Lam, R. W., Frey, B. N., Milev, R., Müller, D. J., Blier, P., Soares, C. N., Parikh, S. V., Turecki, G., Foster, J. A., Rotzinger, S., Kennedy, S. H., & Farzan, F. (2023). Developing an Electroencephalography-Based Model for Predicting Response to Antidepressant Medication. JAMA Network Open, 6(9), e2336094. 10.1001/jamanetworkopen.2023.36094

Sherman, M. A., Lee, S., Law, R., Haegens, S., Thorn, C. A., Hämäläinen, M. S., Moore, C. I., & Jones, S. R. (2016). Neural mechanisms of transient neocortical beta rhythms: Converging evidence from humans, computational modeling, monkeys, and mice. Proceedings of the National Academy of Sciences, 113(33), E4885–E4894. 10.1073/pnas.1604135113

Shin, H., Law, R., Tsutsui, S., Moore, C. I., & Jones, S. R. (2017). The rate of transient beta frequency events predicts behavior across tasks and species. eLife, 6, e29086. 10.7554/eLife.29086

Trivedi, M. H., Chin Fatt, C. R., Jha, M. K., Cooper, C. M., Trombello, J. M., Mason, B. L., Hughes, J., Gadad, B. S., Czysz, A. H., Toll, R. T., Fuller, A. K., Sethuram, S., Mayes, T. L., Minhajuddin, A., Carmody, T., & Greer, T. L. (2020). Comprehensive phenotyping of depression disease trajectory and risk: Rationale and design of Texas Resilience Against Depression study (T-RAD). Journal of Psychiatric Research, 122, 22–32. 10.1016/j.jpsychires.2019.12.004

Trivedi, M. H., McGrath, P. J., Fava, M., Parsey, R. V., Kurian, B. T., Phillips, M. L., Oquendo, M. A., Bruder, G., Pizzagalli, D., Toups, M., Cooper, C., Adams, P., Weyandt, S., Morris, D. W., Grannemann, B. D., Ogden, R. T., Buckner, R., McInnis, M., Kraemer, H. C., … Weissman, M. M. (2016). Establishing moderators and biosignatures of antidepressant response in clinical care (EMBARC): Rationale and design. Journal of Psychiatric Research, 78, 11–23. 10.1016/j.jpsychires.2016.03.001

Trivedi, M. H., South, C., Jha, M. K., Rush, A. J., Cao, J., Kurian, B., Phillips, M., Pizzagalli, D. A., Trombello, J. M., Oquendo, M. A., Cooper, C., Dillon, D. G., Webb, C., Grannemann, B. D., Bruder, G., McGrath, P. J., Parsey, R., Weissman, M., & Fava, M. (2018). A Novel Strategy to Identify Placebo Responders: Prediction Index of Clinical and Biological Markers in the EMBARC Trial. Psychotherapy and Psychosomatics, 87(5), 285–295. 10.1159/000491093

Van Dijk, H., Van Wingen, G., Denys, D., Olbrich, S., Van Ruth, R., & Arns, M. (2022). The two decades brainclinics research archive for insights in neurophysiology (TDBRAIN) database. Scientific Data, 9(1), 333. 10.1038/s41597-022-01409-z

Vaughn, D. A., Marino, B., Engelbertson, A., Dojnov, A., Weiss, N., Vila-Rodriguez, F., Nanos, G., & Downar, J. (2024). Real-world effectiveness of a single-day regimen for transcranial magnetic stimulation using Optimized, Neuroplastogen-Enhanced techniques in Depression (ONE-D). 10.21203/rs.3.rs-5679327/v1

Voetterl, H. T. S., Sack, A. T., Olbrich, S., Stuiver, S., Rouwhorst, R., Prentice, A., Pizzagalli, D. A., Van Der Vinne, N., Van Waarde, J. A., Brunovsky, M., Van Oostrom, I., Reitsma, B., Fekkes, J., Van Dijk, H., & Arns, M. (2023). Alpha peak frequency-based Brainmarker-I as a method to stratify to pharmacotherapy and brain stimulation treatments in depression. Nature Mental Health. 10.1038/s44220-023-00160-7

Warden, D., Rush, A. J., Trivedi, M. H., Fava, M., & Wisniewski, S. R. (2007). The STAR*D project results: A comprehensive review of findings. Current Psychiatry Reports, 9(6), 449–459. 10.1007/s11920-007-0061-3

Webb, C. A., Dillon, D. G., Pechtel, P., Goer, F. K., Murray, L., Huys, Q. J., Fava, M., McGrath, P. J., Weissman, M., Parsey, R., Kurian, B. T., Adams, P., Weyandt, S., Trombello, J. M., Grannemann, B., Cooper, C. M., Deldin, P., Tenke, C., Trivedi, M., … Pizzagalli, D. A. (2016). Neural Correlates of Three Promising Endophenotypes of Depression: Evidence from the EMBARC Study. Neuropsychopharmacology, 41(2), 454–463. 10.1038/npp.2015.165

Williams, L. M., Rush, A. J., Koslow, S. H., Wisniewski, S. R., Cooper, N. J., Nemeroff, C. B., Schatzberg, A. F., & Gordon, E. (2011). International Study to Predict Optimized Treatment for Depression (iSPOT-D), a randomized clinical trial: Rationale and protocol. Trials, 12(1), 4. 10.1186/1745-6215-12-4

Wu, W., Zhang, Y., Jiang, J., Lucas, M. V., Fonzo, G. A., Rolle, C. E., Cooper, C., Chin-Fatt, C., Krepel, N., Cornelssen, C. A., Wright, R., Toll, R. T., Trivedi, H. M., Monuszko, K., Caudle, T. L., Sarhadi, K., Jha, M. K., Trombello, J. M., Deckersbach, T., … Etkin, A. (2020). An electroencephalographic signature predicts antidepressant response in major depression. Nature Biotechnology, 38(4), 439–447. 10.1038/s41587-019-0397-3

Zhdanov, A., Atluri, S., Wong, W., Vaghei, Y., Daskalakis, Z. J., Blumberger, D. M., Frey, B. N., Giacobbe, P., Lam, R. W., Milev, R., Mueller, D. J., Turecki, G., Parikh, S. V., Rotzinger, S., Soares, C. N., Brenner, C. A., Vila-Rodriguez, F., McAndrews, M. P., Kleffner, K., … Farzan, F. (2020). Use of Machine Learning for Predicting Escitalopram Treatment Outcome From Electroencephalography Recordings in Adult Patients With Depression. JAMA Network Open, 3(1), e1918377. 10.1001/jamanetworkopen.2019.18377

